# Assessing the potential impact of transmission during prolonged viral shedding on the effect of lockdown relaxation on COVID-19

**DOI:** 10.1101/2020.06.12.20129213

**Authors:** Burcu Tepekule, Anthony Hauser, Viacheslav N. Kachalov, Sara Andresen, Thomas Scheier, Peter W. Schreiber, Huldrych F. Günthard, Roger D. Kouyos

## Abstract

A key parameter in epidemiological modeling which characterizes the spread of an infectious disease is the mean serial interval. There is increasing evidence supporting a prolonged viral shedding window for COVID-19, but the transmissibility in this phase is unclear. Based on this, we build a model including an additional compartment of infectious individuals who stay infectious for a longer duration than the reported serial interval, but with infectivity reduced to varying degrees. We find that such an assumption also yields a plausible model in explaining the data observed so far, but has different implications for the future predictions in case of a gradual easing on the lockdown measures. Considering the role of modeling in important decisions such as easing lockdown measures and adjusting hospital capacity, we believe that it is critically important to consider a chronically infectious population as an alternative modeling approach to better interpret the transmission dynamics of COVID-19.

## 1 Introduction

Mathematical models have been extensively used to understand the epidemic characteristics of oubreaks, in predicting future outcomes, and in shaping the national responses regarding control measures [1, 2]. Despite the time pressure, a considerable amount of work has been dedicated to modeling the pandemic of novel coronavirus (SARS-CoV-2) infections that began in China in late 2019 [3–6]. Although most of these studies are based on existing epidemic models such as SIR and SEIR-models, several features of the COVID-19 pandemic have been independently explored, leading to different generalizations of similar dynamical models. On one hand, having a variety of models is central to get a notion of the model sensitivity, on the other, it shows that different assumptions are equally favorable to explain the observed data given the right set of parameter choices, whereas they might lead to different projections on how the epidemic would follow in the future [7, 8]. This variability in future projections becomes especially important when a perturbation, such as the imposition or release of the control measures, is introduced to the dynamical system.

A key epidemiologic variable that characterizes the spread of an infectious disease is the mean serial interval [9], i.e., the time between successive cases in a chain of transmission. Li *et al*. [10] *estimated the serial interval distribution to have a mean of 7*.5 (95%CI 5.5 −19) days based on 6 observations, whereas Ganyani *et al*. estimated the serial interval distribution to have a mean of 5.20 (95%CI 3.78 −6.78) days for Singapore and 3.95 (95%CI 3.01 −4.91) days for Tianjin [11], Bi *et al*. estimated the serial interval distribution to have a mean of 6.3 (95%CI 5.2− 7.6) days [12], He *et al*. estimated the serial interval distribution to have a mean of 5.8 (95%CI 4.8 −6.8) days [13], and Hiroshi *et al*. estimated the serial interval distribution to have a mean of 4.7 (95%CI 3.70 −6.00) days. Considering all these studies, infectiousness is estimated to decline quickly within 4 to 8 days on average.

By contrast, certain cases arouse concern about prolonged shedding of SARS-CoV-2 after recovery [14]. Moreover, several studies show proof of active virus replication in upper respiratory tract tissues and prolonged viral shedding even after seroconversion for COVID-19, implying that the contagious period of COVID-19 might last more than one week after clinical recovery in a fraction of patients [15, 16]. De Chang *et al*. reported patients to be virus positive even after the resolution of symptoms up to 8 days [17]. Similarly, Young *et al*. reported a median duration of 12 days for viral shedding [18], and Zhou *et al*. observed a median duration of 20 days [19]. Tan *et al*. reported a special case where the duration of viral shedding persisted for 49 days from illness onset [20]. Considering that the duration of infectiousness is a critical parameter in dynamical models used for predictive purposes, it is important to consider the epidemiological plausibility of a longer shedding window than the reported serial interval in the literature and investigate its impact on model outcomes.

To do so, we first develop a generalized SEIR model by segregating the infectious compartment into two as “primarily infectious” and “chronically infectious” population. We assume that primarily infectious individuals have a higher infectiousnesss within the time window conventially considered as the serial interval, during when they have the potential to develop symptoms and therefore be hospitalized. Afterwards, we assume that the non-hospitalized infecteds transition to the chronically infectious phase before recovery and become less infectious, but may stay infectious for a longer duration. By doing so, we include the possibility of a prolonged viral shedding window in our model. Using the incidence and fatality data from different countries, we first show that our model is also a plausible candidate for explaining the data observed so far for different levels of infectiousness assumed for the chronically infectious population. Based on this conclusion, we explore the model predictions in case of gradual easing on the lockdown measures (relaxation) in Switzerland. Our results show that the model predictions vary in one order of magnitude range for the data considered (daily cases, daily deaths, patients at the hospital ward, and patients at the ICU) for different levels of infectiousness assumed for the chronically infectious population. This variability is especially important when national policies on control measures are being formed, and also for the healthcare systems if projections such as the occupancy of the hospital ward or the ICU are calculated using similar dynamical models.

## 2 Methods

### 2.1 Mathematical Model

To describe the dynamics of the COVID-19 pandemic, we generalize the susceptible-exposed-infected-removed (SEIR) compartmental model by including eight different states denoted by *S*(*t*), *E*(*t*), *I*_*p*_(*t*), *I*_*c*_(*t*), *H*(*t*), *ICU* (*t*), *R*(*t*), and *X*(*t*), representing the number of susceptible individuals, exposed (infected but not yet infectious) individuals, primarily infectious individuals, chronically infectious individuals, hospitalized patients, patients in ICUs, recovered (immune) individuals, and deceased individuals at time *t*, respectively. To model the prolonged viral shedding in case of COVID-19, we segregate the infectious compartment into two by introducing two different compartments, namely the primarily infectious (*I*_*p*_) and the chronically infectious (*I*_*c*_) individuals. After the incubation period is complete, exposed individuals become primarily infectious where they stay infectious within the reported duration of the serial interval of COVID-19. Conventionally, these individuals are assumed to stop being infectious and therefore stop contributing to the disease spread when the serial interval is complete. Our purpose by including another step before recovery, i.e., the chronically infectious compartment, is to model a scenario such that the primarily infectious individuals transition to a state where they are less infectious but they may stay infectious for a longer duration than the serial interval, i.e. continue spreading the infection with reduced transmissibility.

Transitions between different compartments are illustrated in Fig. 1, which can be translated into a system of ordinary differential equations, where each arrow, i.e., each process, is associated with a rate. This system is given by the Eq. set 1, including the rates of processes as model parameters, and describes the rate of change of compartments over time. Model parameters are given in Table 1 with their corresponding descriptions. An additional compartment *C*(*t*) is included in the Eq. set 1 to calculate the cumulative number of the positively diagnosed cases in the community, and does not play a role in the disease dynamics.

**Table 1:** Model parameters given with their descriptions, constrained ranges, and prior distributions.

| Notation | Description | Constained range or definition | Prior distribution <sup>1</sup> |
| --- | --- | --- | --- |
| $R_0^p$ | Basic reproduction number of the primarily infectious population | $0 - \infty$ | $R_0^p \sim \mathcal{N}(2.5, 0.5)$ |
| $r_c$ | Reduction in infectiousness due to being chronic | $0\% - 100\%$ | Fixed to a different value for each simulation. |
| $R_0^c$ | Basic reproduction number of the chronically infectious population | $R_0^c = R_0^p(1 - r_c)$ | Conditioned on $R_0^p$ and $r_c$. |
| $r_L$ | Effect of lockdown in reducing infectiousness | $0\% - 100\%$ | $r_L \sim \beta(1, 1)$ |
| $m_L$ | Slope of reduction in infectiousness due to lockdown | $0.5 - 1.5$ | $m_L \sim 0.5 + \beta(1, 1)$ |
| $s_L$ | Time lag of reduction in infectiousness due to lockdown | $0 - \infty$ | $s_L \sim \exp(1/5)$ |
| $r_{\text{lock}}(t)$ | Time dependent effect of the lockdown on the transmission rate | Given by Eq. 2 | Conditioned on $r_L$, $r_c$, $m_L$, and $s_L$. |
| $1/\tau$ | Incubation period | $0 - \infty$ | $\tau \sim \exp(1/2.5)$ |
| $1/\gamma_p$ | Duration of infection of $I_p$ | $0 - \infty$ | $\gamma_p \sim \exp(1/2.5)$ |
| $1/\gamma_c$ | Duration of infection of $I_c$ | 14 days | Fixed for all simulations. |
| $\beta_p$ | Transmission rate of $I_p$ | Given by Eq. 3 | Conditioned on $r_{\text{lock}}(t)$, $R_0$, and $\gamma_p$. |
| $\beta_c$ | Transmission rate of $I_c$ | Given by Eq. 4 | Conditioned on $r_{\text{lock}}(t)$, $R_0$, $\gamma_c$, and $r_c$. |
| $1/\gamma_H$ | Duration of hospital ward stay | $0 - \infty$ | $\gamma_H \sim \exp(1/12)$ |
| $1/\gamma_{ICU}$ | Duration of ICU stay | $0 - \infty$ | $\gamma_{ICU} \sim \exp(1/12)$ |
| $\epsilon_H$ | Rate of direct H admission | $0 - \infty$ | $\epsilon_H \sim \mathcal{N}(0.08, 0.02)$ |
| $\epsilon_{H2I}$ | Transfer rate from H to ICU | $0 - \infty$ | $\epsilon_{H2I} \sim \mathcal{N}(0.4, 0.08)$ |
| $\epsilon_x$ | Death rate from ICU | $0 - \infty$ | $\epsilon_x \sim \mathcal{N}(0.4, 0.08)$ |
| $r_d^p$ | Diagnosis rate of $I_p$ | $0 - \infty$ | $r_d^p \sim \mathcal{N}(0.2, 0.03)$ |
| $r_d^c$ | Diagnosis rate of $I_c$ | $0 - \infty$ | $r_d^c \sim \mathcal{N}(0.075, 0.015)$ |
| $R_0$ | Total basic reproduction number | $R_0 = R_0^p + (1 - \epsilon_H)R_0^c$ | Conditioned on $R_0^p$, $R_0^c$ and $\epsilon_H$. |
| $E(0)$ | Initial frequency of the exposed compartment | $0\% - 100\%$ | $r_d^c \sim \beta(1, 10^3)$ |
| $S(0)$ | Initial frequency of the susceptible compartment | $1 - E(0)$ <sup>2</sup> | Conditioned on $E(0)$ |
| $N$ | Population size | — | Fixed specific to the country used for fitting. |

**Figure 1:**
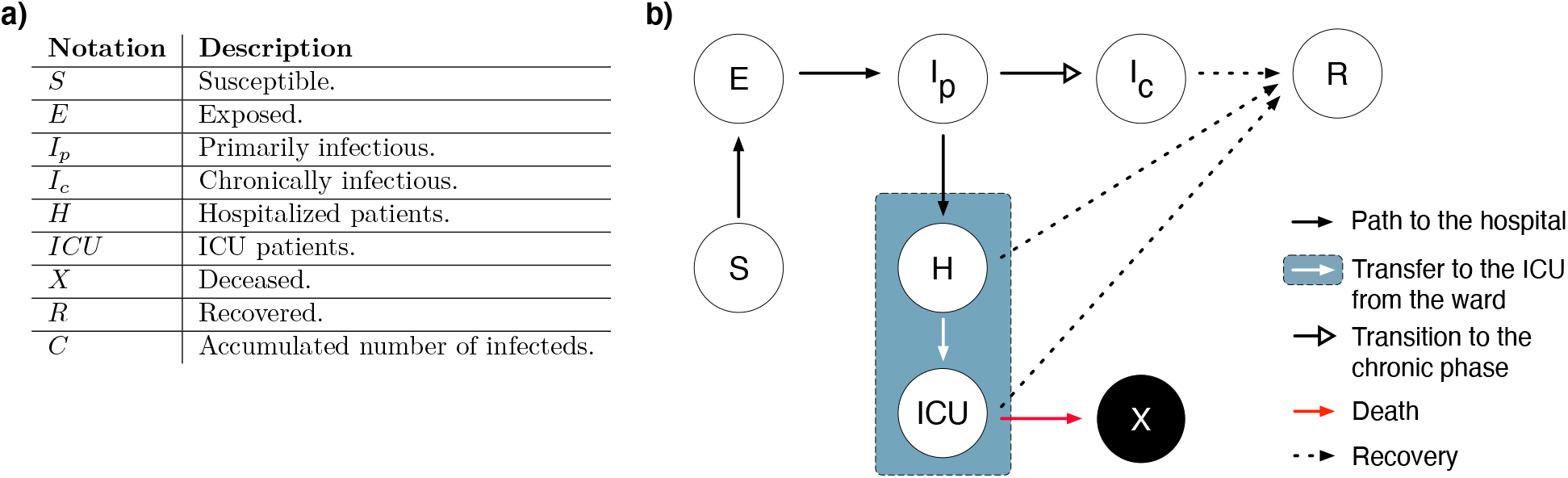
**a)** Notation of the compartments and their corresponding descriptions. **b)** Schematic of the dynamical model given by Eq. set 1.

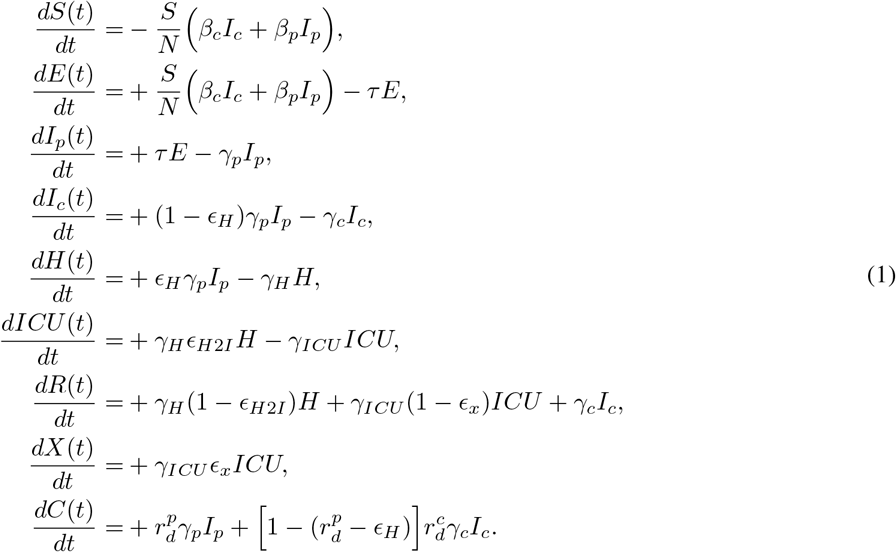

Time-dependent decrease in the transmission of SARS-CoV-2 due to lockdown measures is modeled by a sigmoid function [21], and denoted by *r*_*lock*_(*t*), such that

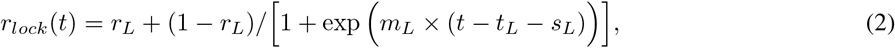

where *r*_*L*_, *t*_*L*_, *m*_*L*_, and *s*_*L*_ denote the final effect of the lockdown, start date of the lockdown, slope of the decrease in transmissibility, and the time delay between implementation and effect of the lockdown, respectively. *r*_*lock*_(*t*) is used as a multiplicative factor in modeling the transmission rate in a time-dependent manner.

The reduced transmissibility of *I*_*c*_ is modeled via including a reduction coefficient *r*_*c*_ as a multiplicative factor to its transmission rate, resulting in two different transmission rates *β*_*p*_ and *β*_*c*_ for *I*_*p*_ and *I*_*c*_ compartments, such that,

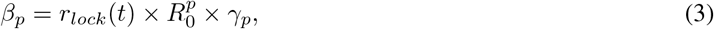

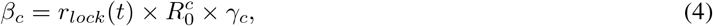

Where 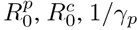, and 1/*γ*_*c*_ denotes the basic reproduction number of the primarily infectious population, the basic reproduction number of the chronically infectious population, duration of primarily infectious phase, and the duration of chronically infectious phase, respectively. We assume that individuals who develop symptoms do so only during the primarily infectious phase, and therefore hospitalization is only possible before they transition to the chronically infectious phase. We do not assume any a priori information regarding the testing policy, therefore a positive diagnosis is possible for both primarily and chronically infected individuals, and they contribute to the cumulative number of the positively diagnosed cases with the rates 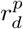 and 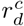, respectively.

### 2.2 Model Fitting and Parameter Estimation

We implemented two stages of model fitting. The first stage aims to compare the predictions of the model given by Eqn. set 1 for different values of ≤ *r*_*c*_, where 0%≤ *r*_*c*_ ≤ 100% with a step size of 2% to cover different scenarios with different levels of chronic infectiousness. We refer to the model with *r*_*c*_ = 100% as the “null model”, and assume that the primarily infectious individuals do not have a prolonged viral shedding, but they still can be diagnosed during the chronic phase, meaning that their test results can still be positive although they are not infectious. We then fit the model simultaneously to the data on the number of daily confirmed cases and the number of daily deaths for various countries, reported by the World Health Organization (WHO) [22]. The deviations between the model output and the data are assumed to follow a Negative Binomial distribution. Dispersion parameters of the Negative Binomial distributions are estimated seperately for both the number of daily confirmed cases and the number of daily deaths during model fitting.

When fitting the model, we fixed the reduction in infectiousness parameter *r*_*c*_ to different values varying between 0% to 100%. Duration of infectiousness of the *I*_*c*_ compartment is fixed to 14 days for all simulations. Other parameters are allowed to vary within their respective ranges, given in Table 1.

During model fitting, we leave out a certain amount of datapoints and use them later to calculate the prediction error, which is defined as the mean Euclidian distance between the model prediction and the datapoints that are not used for fitting. To interpret the prediction error of the models with prolonged viral shedding (0%≤ *r*_*c*_ ≤ 98%) relative to the null model (*r*_*c*_ = 100%), we normalize each prediction error value by the prediction error of the null model for a given dataset size, and call it the relative prediction error (RPE). By definition, the RPE of the null model is 1. Any RPE value below 1 indicates that the corresponding model has a better predictive power relative to the null model.

All datapoints before the introduction of the lockdown measures are included in the fitting procedure to have a better estimate of the effect of the lockdown. Due to the uncertainty of the quantitative effects of the easing on the lockdown measures (relaxation), datapoints after the relaxation are excluded from RPE calculations. By comparing the RPE values of the models with and without the prolonged viral shedding, we aim to demonstrate that both types of model yield to a certain predictive power that makes it difficult to reject the possibility of the existence of a chronically infectious population from a modeling perspective.

Building upon this conclusion, the second stage of the model fitting demonstrates how the impact relaxation would differ given different levels of reduction in infectiousness for the *I*_*c*_ compartment. We use the data provided by [23] for Switzerland, and fit the model simultaneously to four datasets : the number of daily confirmed cases, the number of daily deaths, the number of patients at the hospital ward at a given day, and the number of patients at the ICU at a given day. Only Swiss data is used to asses the impact of relaxation because it is the only country to our knowledge where data on both the hospitalized and the ICU patients are publicly available in addition to the number of daily confirmed cases and the number of daily deaths. This enabled us to quantify the effects of relaxation on the capacity requirements of healthcare systems. On the other hand, we chose to exclude Swiss data from the first stage of model fitting due to its short lockdown duration relative to other countries included in that analysis.

Using the parameters we obtained via fitting, we predict the outcomes of a gradual relaxation scenario both for the null model and the model including the chronically infectious population with different *r*_*c*_ values. Relaxation is modeled as an increase in transmissibility, and characterized as a sigmoid function. It is similar to the time-dependent effect of the lockdown (*r*_*lock*_(*t*)) given by Eqn. 2, such that

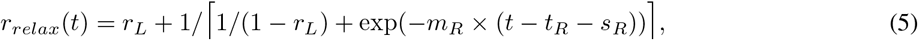

where *t*_*R*_, *m*_*R*_, and *s*_*R*_ denote the start date of the relaxation (27^*th*^ of April for Switzerland, 63 days after the first case is observed), slope of increase in transmissibility (set to 0.1 for a gradual relaxation), and the time delay until the effect of the relaxation takes place (set to 60 days for a gradual relaxation), respectively. *r*_*relax*_(*t*) is used as a multiplicative factor in a similar fashion to *r*_*lock*_(*t*). Note that the parametrization of *r*_*relax*_(*t*) does not depend on data, and its parameter values are chosen such that it demonstrates a gradual increase in transmissibility over the months following the start of relaxation.

For both stages of fitting, we implemented the model in a Bayesian framework using Stan [24]. Prior distributions of the parameters used during fitting are given in Table 1. All code and data are available from https://github.com/burcutepekule/covid_prolonged_shedding.

## 3. Results

### 3.1 Possibility of a chronically infectious population

To have as many data points as possible for relative prediction error (RPE) calculations, we have chosen the countries with the longest durations between the start of the lockdown measures and the relaxation, which are Italy, France, Spain, Greece, province of Hubei, and the U.S.A. During model fitting, we used all the datapoints until the start of the lockdown with an additional 5 days of observation, and gradually increased the number of datapoints (the dataset size) by another 5 days to systematically explore the effects of the dataset size on the prediction error.

We find that both the null model and the models with prolonged viral shedding provide almost identical fits for the observed data independent of the dataset size, however they differ in their predictive abilities. For Italy, the models with a higher level of infectiousness (lower *r*_*c*_) provide a better prediction relative to the null model (*r*_*c*_ = 100%) for both the number of confirmed cases and deaths when the size of the dataset used for fitting is small (Figs. 2 **a), b), e)**, and **f)**). As the dataset size used for fitting increases, both the null model and the models with prolonged viral shedding provide similar short-term predictions and narrower confidence intervals (Figs. 2 **c), d)**). In case of Greece, we observe that all RPE values for both the number of daily deaths and daily cases are close to 1 (SI Fig. 1), meaning that there is not a substantial difference in the predictive capacity of the null model relative to the models including prolonged viral shedding with different levels of infectiousness. For Spain, RPE values decrease as the level of infectiousness decreases (*r*_*c*_ increases) for both the number of daily cases and daily deaths, and the models with prolonged viral shedding provide a better prediction for smaller datasets when the level of infectiousness is below 20% (*r*_*c*_ > 80%). Additionally, using smaller datasets for fitting results in a very poor predictive capacity and very wide confidence intervals for all models including the null model (SI Fig. 2 **a)** and **b)**). Analysis for the U.S.A (SI Fig. 4) and France (SI Fig. 5) provide similar results as Spain. For Hubei, all models perform poorly to predict the number of daily cases (SI Fig. 3 **a)** and **c)**), whereas the models with prolonged viral shedding with higher levels of infectiousness provide better predictions as the dataset size used for fitting increases.

**Figure 2:**
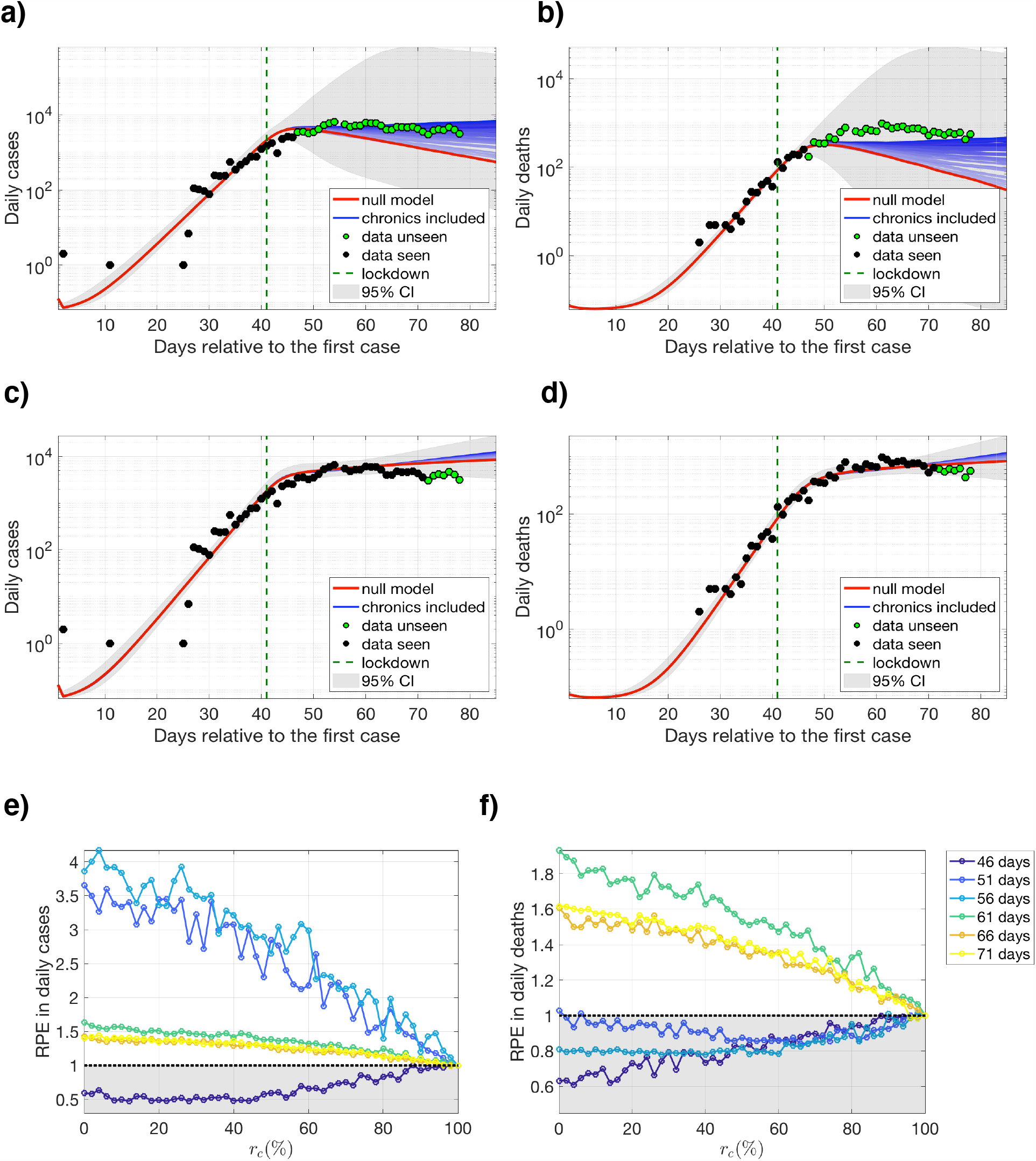
Fitting, prediction, and relative prediction error (RPE) results for Italy, calculated using different levels of infectiousness for the chronically infectious population and different sizes of datasets. Model outcomes for the number of daily confimed cases (**a), c)**) and daily deaths (**b), d)**) using an additional 5 and 30 days of observed data for model fitting, respectively. Predictions drawn in darker shades of blue represent the fitting results with increased infectiousness of the chronically infectious population, i.e., lower *r*_*c*_ values, and the predictions for the null model (*r*_*c*_ = 100%) are drawn in red. Data points that are used for fitting are drawn in black whereas the datapoints that are excluded from fitting but used for prediction error calculations are drawn in green. Gray areas around the model outcomes represent the union of the confidence intervals calculated for different levels of infectiousness. Relative prediction error (RPE) **e)** for the number of daily cases and **f)** the number of daily deaths for a given *r*_*c*_ value and a given dataset size used for fitting, where *r*_*c*_ = 100% represents the results for the null model, and values below 1 are shaded in gray for a better visualization.

**Figure 3:**
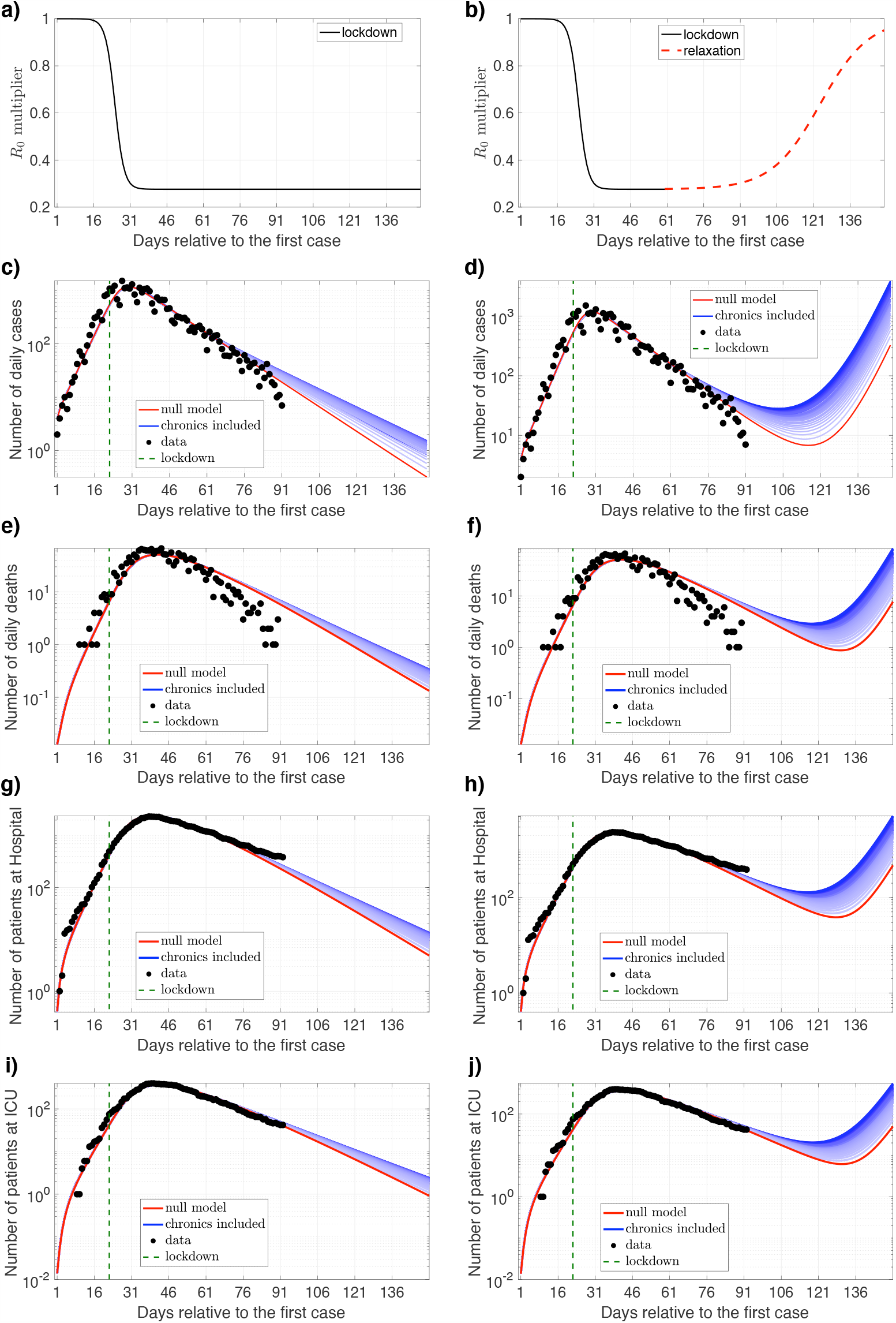
Fitting results and predictions for Switzerland, calculated using different levels of infectiousness for the chronically infectious population, (**a), c), e), g)**, and **i)**) assuming the lockdown measures continue, and (**b), d), f), h)**, and **j)**) assuming a gradual easing on the lockdown measures (relaxation). Time dependent effects of the lockdown and the relaxation are illustrated in **a)** and **b)**, respectively. Predictions drawn in darker shades of blue represent the fitting results with increased infectiousness of the chronically infectious population, i.e., lower *r*_*c*_ values, and the predictions for the null model (*r*_*c*_ = 100%) are drawn in red.

**Figure 4:**
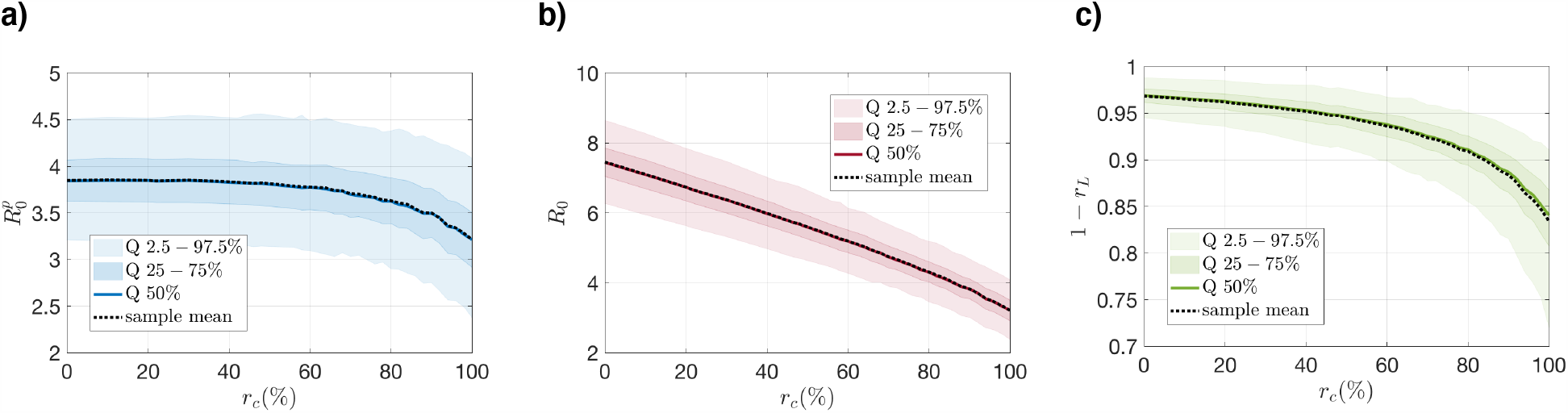
Posterior distributions of the **a)** basic reproduction number of the primarily infectious population 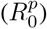, **b)** total basic reproduction number 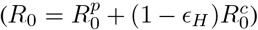, and **c)** the final reduction in infectiousness due to lockdown for a given *r*_*c*_ value.

As the dataset size used for fitting increases, both the null model and the models with prolonged viral shedding provide similar short-term predictions (Panels **c)** and **d)** for SI Figs. 1-5 and Fig 2.). This is to be expected since the data points used for fitting can be explained equally well using all models, and the differences in infectiousness level (*r*_*c*_) manifest itself more as the prediction horizon increases. Consequently, RPE curves with larger dataset sizes result in lower RPE values in general for all countries.

### 3.2 Impact of relaxation

Data for Switzerland is used to simulate the impact of a gradual easing on the lockdown measures (relaxation) both for the null model and the model with prolonged viral shedding for different levels of reduction in infectiousness (different values of *r*_*c*_).

We observe that the predictions for both the continuation of the lockdown and the relaxation present similar shapes, but the quantitative difference varies by one order of magnitude range for different *r*_*c*_ values (Fig. 3). In case of the continuation of the lockdown, a longer duration is required for the number of daily deaths and daily cases to go down to the same value as the infectiousness of the chronically infectious population increases (Figs. 3 **c), e), g)**, and **i)**). In case of relaxation, both the number of daily cases and the number of daily deaths start to increase faster as the infectiousness of the chronically infectious population increases, and reach almost a ten fold higher value within two weeks following the start of the relaxation (Figs. 3 **d)** and **f)**). This increase is also reflected in the number of patients observed in the hospital ward and the ICU (Figs. 3 **h)** and **j)**), indicating a faster influx to the health facilities under the assumption of a highly chronically infectious population.

The fact that observed data can be explained equally well by all *r*_*c*_ values varying from 0% to 100% (null model) is partially due to the flexibility of the fitting procedure, which allows other parameters to be adjusted for a given *r*_*c*_ value. Most parameters are free to vary, but their prior distributions are informed such that the hyperparameters (parameters of the prior distributions) align with the reported values in the literature (Table 1). As an example, both the incubation period (1/*τ*) and the duration of infectiousness of the primarily infectious population (1/*γ*_*p*_) have the mean of 2.5 days, resulting in a serial interval distribution with a mean of 5 days, in agreement with the reported values in the literature for COVID-19 (see Introduction). Similarly, the basic reproduction number of the primarily infectious population 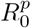 is normally distributed with a mean of 2.5, which is the average value reported for basic reproduction number of COVID-19 in many countries [10, 25]. Mean values of the prior distributions of the parameters related to hospitalization (*γ*_*H*_, *γ*_*ICU*_, *ϵ*_*H*_, *ϵ*_*H*_2_*I*_, and *ϵ*_*x*_) are adopted from Ferguson *et al*. [26] and Verity *et al*. [27], and given a variance such that they can be adjusted specifically for each country during the fitting procedure.

The *r*_*c*_ dependent posterior distributions for 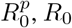, *R*_0_, and *r*_*L*_ provide a good example to demonstrate the flexibility of the fitting procedure (Fig 4). As expected, the final reduction in infectiousness due to lockdown (1 − *r*_*L*_), basic reproduction number of the primarily infectious population 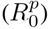, and the total basic reproduction number 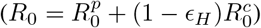 is estimated to be lower as the infectiousness of the chronically infectious population decreases (as *r*_*c*_ increases) to explain the observed data.

Although the models we have explored so far assume different levels of infectiousness during the prolonged viral shedding window including no infectivity at all (null model), they all assume that the infected individuals are tested and positively diagnosed with a certain rate during the chronically infectious phase. This is not a common assumption in other modeling studies regarding COVID-19. For comparison, we also applied both stages of model fitting using a chronic-free model where the *I*_*c*_ compartment is completely omitted. We find that the chronic-free model substantially underpredicts both the number of daily cases and the number of daily deaths when the size of the dataset used for fitting is small (SI Fig. 6). Similar to the models explored so far, its predictive capacity increases as the dataset size used for fitting increases. A chronic-free model can also explain the observed data equally well (SI Figs. 6 and 7), but this is only possible with a considerably lower basic reproduction number combined with a considerably lower lockdown effect compared to the models including the *I*_*c*_ compartment (SI Fig. 8).

## 4 Discussion

The model presented in this work explores the epidemiological plausibility of a prolonged viral shedding window for the COVID-19 pandemic, and quantifies the impact of a gradual easing on the lockdown measures (relaxation) given different assumptions on the infectiousness of a chronically infectious population.

Our results show that having a chronically infectious population, i.e., individuals that are less infectious but infectious for a longer duration, is not a possibility that can be easily rejected from an epidemiological perspective. This conclusion is based on two main results. First, the data that has been observed so far can be explained equally well by the model with prolonged viral shedding for a variety of different levels of reduced infectiousness as well as the null model, i.e., the model without prolonged viral shedding. Although this is partially due to the flexibility of the fitting procedure, the choice of hyperparameters (parameters of the prior distributions) indicate that all fits for a given reduced infectiousness value are possible for a set of reasonable model parameters, and therefore as favorable as the null model from a modeling perspective.

Second, it is not clear whether the null model or the model with prolonged viral shedding provides more accurate predictions for the prospective data that is not included in the fitting procedure. The uncertainty around the prediction error values for different countries, different types of data (confirmed cases or deaths), and different data sizes used for fitting indicates that it is not always the null model that has the higher predictive power. Consequently, our analysis shows that it is not possible to either accept or reject the existence of a chronically infectious population with reasonable certainty from a modeling perspective.

The fact that observed data can also be explained with a model including prolonged viral shedding raises certain questions about the interpretation of the epidemic curve, acquired immunity, and the current testing policies. Assuming a relatively short serial interval for a model that does not consider a prolonged viral shedding window results in more optimistic projections about epidemic control, as clearly demonstrated in Figs. 2 and 3. Countries that were very successful in their initial control measures and therefore experienced a very steep decline in the number of daily confirmed cases might choose to ease the control measures too soon. We still lack a full understanding of the viral shedding window of COVID-19, and therefore might have a biased opinion on the number of infectious individuals in the community. This once again emphasizes the infectiousness of COVID-19 and the significance of frequent testing although the number of confirmed cases are in decline.

Building up on these conclusions and concerns, we investigate how the population dynamics would follow in case of a gradual easing on the lockdown measures (relaxation) in Switzerland considering different levels of infectiousness during a prolonged viral shedding window. Our results show that although the predictions present a similar shape for different levels of reduced infectiousness for the chronically infectious population, the quantitative difference varies by one order of magnitude range for all four signals (daily cases, daily deaths, patients at the hospital ward, and patients at the ICU). This variability is especially important for the healthcare systems if projections such as the occupancy of the hospital ward or the ICU are calculated using similar dynamical models.

Using simplified compartmental models such as the one in this study has certain limitations. First, it does not consider the stochastic effects that the system is subject to, which become more important as the number of infecteds decrease in the community. Second, it assumes a well-mixed population, and does not consider the contact structure and the demographic information which are both relevant to the disease spread. Nevertheless, we believe that these two drawbacks of our modeling approach influence the models with and without the prolonged viral shedding to a similar degree, if not penalizing the model with prolonged viral shedding for producing more pessimistic projections since the number of infecteds will be higher in frequency relative to the null model.

It is still debated whether the patients who recover from COVID-19 and test positive for the virus after their recovery are still infectious or not. Nevertheless, it is clear that these positive test results contribute to the data on the number of daily confirmed cases. However, current modeling studies regarding COVID-19 neglect this fact and assume that all positive test results are recorded within the duration of the serial interval only. Our results show that this assumption might lead to an underestimation of both the reproduction number and the effect of the lockdown, leading to a potential underprediction for the prospective data.

In conclusion, It is not possible to either prove or disprove the existence of a certain compartment of individuals purely by modeling. Including a chronically infectious population in our model was motivated by the evidence reported for prolonged viral shedding in the literature [14–20], and attempted to test whether this is also a plausible modeling approach to explain the data observed so far. Given that different assumptions on the infectiousness level during a prolonged viral shedding window can result in similar descriptions of the observed data in retrospect but different outcomes in prospect, it is important to consider a chronically infectious population from a modeling perspective when national policies are being imposed.

## Data Availability

All code and data are available from https://github.com/burcutepekule/covid_prolonged_shedding.

## 5 Acknowledgement

We gratefully acknowledge Dr. Julien Riou for his valuable comments and discussions.

## Footnotes

1 𝒩, *β*, exp denotes the Normal, Beta, and Exponential distributions respectively.

2 All other compartments (*I*_*p*_, *I*_*c*_, *H, ICU, R*, and *X*) are assumed to be zero at *t* = 0, and the first case is assumed to be observed at *t* = 1.

